# Malaria vaccination coverage, completeness, and timeliness among children aged 6–24 months: a cross-sectional survey conducted in the Western region of Cameroon, 1 year following the vaccine introduction

**DOI:** 10.1101/2025.09.24.25336598

**Authors:** Jerome Ateudjieu, Merveille Claire Nana Djapou, Benjamin Kevin Bekoa Onana, Abdias Aron Tatsabong Tiomeni, Gretta Ludivine Okoumokath Mpande, Donald Kapso Nanguep, Collins Buh Nkum, Dora Winny Ateudjieu Kenfack, Anne Cecile Bissek

## Abstract

Malaria vaccination was introduced as part of Cameroon EPI vaccine to prevent malaria morbidity and mortality among children aged 6-24 months. This study aims to assess the coverage, completeness, and timeliness of this vaccine, and explore parental perceptions on children access to vaccination. This was a community-based, descriptive cross-sectional study targeting children aged from 6 to 24months and caregivers, selected by stratified cluster random sampling in Foumbot and Foumban health districts. Data were collected from caregivers by trained and supervised enumerators using a face-to-face, pretested questionnaire. Malaria vaccine coverage, completeness, and timeliness were estimated with 95% confidence interval. The contribution of caregivers’ perception on the first dose malaria vaccination status was explored by estimating crude Odd Ratio (cOR) and Adjusted Odd ratio (aOR) estimated from mixed effect logistic regression. Of the 55 targeted and reached clusters 399 children were included in this study. Vaccination coverage of the first, second and the third dose malaria vaccination was 31.20% (95%CI 30.38 - 32.02), 22.61% (95%CI 21.85 - 23.37) and 17.70% (95%CI 16.88 - 18.52) respectively. Among the children exposed to the first dose, 56.49% (95%CI 44.61 - 68.37) completed the third dose. The timeliness of the three administered doses was around 50-57%. In multivariate analysis, parental perception remained significantly associated with dose 1 malaria administration (aOR 15.54 95%CI 12.63 - 19.10 p= <0.001), dose 1 timeliness (aOR 2.36 95%CI 1.52 - 3.65 p= <0.001) and dose 2 completeness (aOR 9.81 95%CI 7.78 - 12.36 p= <0.001). More than a year after the malaria vaccine introduction in the health districts of Foumbot and Foumban, the performance indicators for this vaccination are below expectations for the vaccine to have a significant impact on reducing malaria morbidity in children. Communicating to positively influence caregivers’ perceptions on malaria vaccination expected to contribute in improving the situation.

## Background

Malaria remains a public health challenge worldwide with the highest burden in Africa [1, 2]. Many interventions have been prioritized to reduce the disease burden including environmental interventions, the use of long-lasting mosquito nets, prophylactic treatment regarding vulnerable groups including pregnant women and children in malaria epidemic areas [3-6]. The impact of these interventions have been notable with significant reduction of disease morbidity and mortality but more needs to be done to control the disease [7]. In line with this, vaccine development has delivered the marketing of RTS,S/AS01, R21/Matrix-M and PfSPZ, with RTS,S/AS01 and R21/Matrix-M recommended by WHO [8-15].

Available literature documents the efficacy, safety and benefits of malaria vaccination in a variety of contexts [7, 11, 16]. These data, consolidated after the pilot projects carried out as part of the Expanded Immunization Programs in Kenya, Ghana and Malawi, and the WHO recommendations that followed, have helped to justify the extension of interventions to other countries, including Cameroon [15-18]. This was followed by important reported challenges that included logistic difficulties, funding constraints, the complexity of administering of the four-dose vaccine, and poor caregivers’ adherence in some contexts [19, 20]. In Cameroon RTS,S/AS01 was introduced in 2024 as a part of expanded national immunization program targeting children aged 6-24 months of 42 health districts with highest malaria burden in children. By the ten month from the beginning of the introduction, administrative data reported 44.8% third dose coverage [21]. Exploring this coverage and caregivers’ perceptions of malaria vaccination in households should generate evidence for planning interventions to reach the maximum number of targeted children.

## Methodology

### 1. Study design

This was a community-based descriptive cross-sectional study targeting children aged 6 to 24 months, selected by stratified cluster random sampling. Data were collected using a face-to-face questionnaire administered to caregivers and a grid to review children vaccination booklet. Malaria vaccine coverage, completeness and timeliness were estimated from the collected data.

### 2. Study setting

The study was conducted in the Foumban and Foumbot health district in the West Region of Cameroon (Figure 1) which were the only two districts in the region benefiting from the malaria EPI vaccination program at the time of the study.

**Figure 1:**
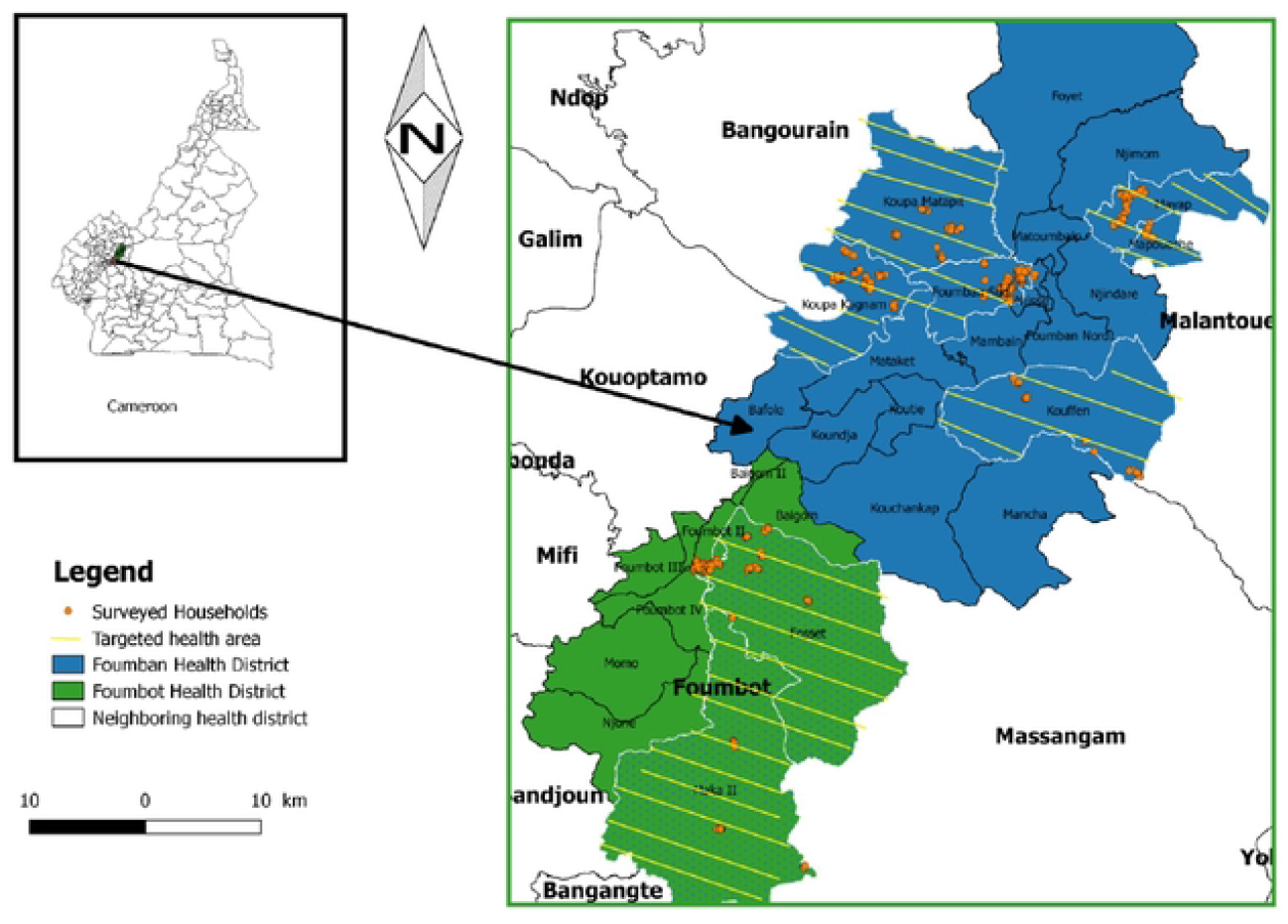
Foumban and Foumbot Districts Map.

### 3. Study population

Children aged 6 to 24 months living in the Foumbot or Foumban health district were eligible. Those with caregivers consenting to participate were included. Children with no caregiver to provide data on vaccination status and those living in health areas with recent history of insecurity were excluded.

### 4. Sample size estimation

The minimum sample size estimated was 392 couple mother-child. This estimation assumed a third dose malaria coverage of 50%, a 95% confidence level, a precision of 7% and a cluster design effect of 2. Assuming to reach about 7 children per cluster, the estimated sample was allocated across 55 clusters.

### 5. Sampling process

Stratified by the population size of Foumban and Foumbot health districts, six and three health areas, respectively, were randomly selected to be part of the study. Each health area, the list of communities and their respective population size was obtained. Clusters were then allocated proportionally to communities’ population size using a stratified systematic sampling approach. In targeted community participants of each cluster were selected by randomly selecting and a direction of recruitment. Households bordering the street were visited following the direction to identify and enrol study participants. Recruitment was considered completed in each cluster when about 7 children were included.

### 6. Data collection tools and variable

Data were collected using a face-to-face administered questionnaire and a grid to review vaccination cards. These tools were adapted from an existing questionnaire that was originally developed to assess documented and undocumented EPI vaccination coverage, timeliness and completeness [22, 23]. The main data collected included the sociodemographic characteristics of the mother and children; child’s malaria vaccination status; caregiver’s exposure to malaria vaccination information, as well as their perception of the malaria vaccination benefits and safety.

### 7. Data collection process

The process involved obtaining informed consent from caregivers, administering the questionnaire and collecting data vaccination data from the vaccination card when accessible.

### 8. Data analysis

We estimated the documented vaccine coverage, timeliness, completeness using numerators and denominators as presented in table 1. Weighting was applied for these estimates considering stratified distribution of allocated clusters per health area. We explored the contribution of caregivers’ perception regarding malaria vaccine on first malaria documented dose coverage and second dose malaria vaccine completeness using case control approach estimating and adjusting Odds Ratio. Adjustment was done using mixed effect logistic regression with confounders identified from the comparison of key characteristics between cases and controls. Controls being children documented vaccinated and cases being children with no documented vaccination. Data were analyzed using SPSS version 24.0 and Excel 2013. Estimates were reported with 95% confidence intervals.

**Table 1:** Definition of denominators and numerators for the estimate of the documented vaccine coverage, timeliness, completeness of the malaria vaccines uptake.

| Estimated indicator | Numerator | Denominator |
| --- | --- | --- |
| Documented coverage first dose malaria vaccine dose | Number of children documented to have received the first malaria vaccine dose | Number of children aged 7-24 months |
| Documented coverage second dose malaria vaccine dose | Number of children documented to have received the second malaria vaccine dose | Number of children aged 8-24 months |
| Documented coverage third dose malaria vaccine dose | Number of children documented to have received the third malaria vaccine dose | Number of children aged 12-24 months of age |
| Timeliness first malaria dose | Number of children documented to have received the first malaria vaccine dose before the end of 7<sup>th</sup> months of age | Number of children vaccinated with first dose malaria vaccine |
| Timeliness second malaria dose | Number of children documented to have received the second malaria vaccine dose before the end of 8<sup>th</sup> months of age | Number of children vaccinated with second dose malaria vaccine |
| Timeliness third malaria dose | Number of children documented to have received the third malaria vaccine dose before 11<sup>th</sup> months of age | Number of children vaccinated with third dose malaria vaccine |
| Two doses completeness among those exposed to 01 dose | Number of children aged 8-24 months (documented) that have received two doses of malaria vaccine | Number of children 8-24 months (documented) that received the first dose of malaria vaccine |
| Two doses completeness among children reached | Number of children aged 8-24 months (documented) that have received two doses of malaria vaccine | Number of children aged 8-24 months |
| Completeness third dose | Number of children aged 12-24 months (documented) that have received three doses of malaria vaccine | Number of children aged 12-24 months (documented) that received the first dose of malaria vaccine |

### 9. Ethical consideration

The present study was conducted to generate evidence for the improvement of malaria vaccination in Cameroon. Its implementation involved household-based data collection from caregiver with risk of increasing access to participants confidential data and compromising their autonomy. Participants were informed of the aims and procedures of the study and were included only when they consented to participate by signing the consent form. Participants’ confidentiality was protected by not collecting identifiable data, password protecting access to collected data and limiting access to dataset to the team in charge of data management and analyses. The protocol was evaluated by the competent ethical review committee (West Region Ethics Review Committee of Human Health Research of Cameroon) and approved with reference number N°/580/29/05/2024/CE/CERSH-OU.

## Results

### 1. Survey coverage

We reached the 55 targeted clusters, and 399 participants all of whom consented to participate with average cluster coverage of 93.41% [95%CI: (91.32 – 95.50)].

### 2. Socio-demographic characteristics of Caregiver and children

The mean age of the caregiver and children were 27.38 ± 7.45 years and 14.24 ± 5.41 months respectively. Socio-demographic characteristics of the study population are presented in table 2. The majority of caregivers (97.1%) were female; 11.5% were adolescents, and 37.5% had a low level of education. Table 3 presents the sociodemographic characteristics of the children.

**Table 2:** Socio-demographic characteristics of caregiver.

| Variable | Modality | Frequency | Percentage(%) |
| --- | --- | --- | --- |
| Sex | Male | 12 | 2.9 |
|  | Female | 387 | 97.1 |
| Age (years) | ≤ 19 | 45 | 11.5 |
|  | 20–49 | 347 | 87 |
|  | 50 and above | 6 | 1.5 |
| Caregiver's relationship with the child | Mother | 373 | 93.8 |
|  | Father | 11 | 2.7 |
|  | Grandmother | 7 | 1.7 |
|  | Older brother or sister | 5 | 1.1 |
|  | Member of the parents' family | 3 | 0.8 |
| Level of education | No school | 19 | 4.5 |
|  | Primary | 129 | 33.0 |
|  | Combined low level of education (No school and primary) | 148 | 37.5 |
|  | Secondary | 242 | 60.2 |
|  | University | 9 | 2.3 |
| Occupation | Housekeeper | 199 | 50.0 |
|  | Farmer | 73 | 17.4 |
|  | Civil servant | 12 | 2.1 |
|  | Entrepreneur | 10 | 2.6 |
|  | Shopkeeper | 24 | 6.1 |
|  | Dressmaker | 47 | 11.8 |
|  | Other | 34 | 10.1 |
| Religion | Christian | 79 | 19.3 |
|  | Muslim | 319 | 80.4 |
|  | Animist | 1 | 0.3 |
| Flour type of household inhabited by the caregiver | Tiles | 57 | 14.6 |
|  | Cemented | 208 | 50.1 |
|  | Carpet | 5 | 1.3 |
|  | Clay | 139 | 34.0 |
| Average number of people in the household of the caregiver | 6.06 ± 2.71 95% CI = [6.013; 6.107] |  |  |

**Table 3:** Socio-demographic characteristics of children.

| Variables | Modalities | Effectives | Percentage |
| --- | --- | --- | --- |
| Sex | Male | 190 | 47.6 |
|  | Female | 209 | 52.4 |
| Age (months) | 6-11 | 135 | 34.1 |
|  | 12-24 | 264 | 65.9 |

### 3. Vaccine coverage, completeness and timeliness

Of the 399 children reached, 208(51.9%) were reported by caregiver to have receive at least 01 dose of malaria vaccination; while 121(29.8%) were documented to have received at least 01 dose of malaria vaccination.

Children vaccination coverage, timeliness and completeness are presented in table 4. Of the included children, 48 (17.70%) were documented vaccinated with the third dose of malaria vaccine. Forty-six children (37.98%) of the 119 children exposed to the first dose completed the third dose. The timeliness of the three doses varied between 50.34% and 56.66%.

**Table 4:** Malaria vaccine coverage, completeness and timeliness.

|  | Frequency | Percentage | 95% CI |
| --- | --- | --- | --- |
| **Documented coverage per dose** |  |  |  |
| Dose 1 (n=372 children aged 7-24) | 119 | 31.20 | 30.38 - 32.02 |
| Dose 2 (n=348 children aged 8-24) | 80 | 22.61 | 21.85 – 23.37 |
| Dose 3 (n=264 children aged 12-24 months) | 48 | 17.70 | 16.88 – 18.52 |
| **Completeness** |  |  |  |
| Two doses completeness among those exposed to 01 dose (n=102 children aged 8-24) | 77 | 74.76 | 66.33 – 83.19 |
| Two doses completeness among children reached (n=348 children aged 8-24) | 77 | 21.65 | 16.72 – 25.28 |
| Three doses completeness among those exposed to 01 dose (n=67 children aged 12-24) | 39 | 56.49 | 44.61 – 68.37 |
| **Timeliness** |  |  |  |
| 1<sup>st</sup> dose (n = 119) | 66 | 55.24 | 53.6 – 56.8 |
| 2<sup>nd</sup> dose (n = 80) | 40 | 50.35 | 48.4 – 52.3 |
| 3<sup>rd</sup> dose (n = 48) | 27 | 56.66 | 48.1– 53.2 |

### 4. Caregivers’ perception on malaria vaccine

Caregivers’ perceptions of the malaria vaccine are presented in Table 5. Of the 399 included caregivers, 325 (81.8%) had positive perceptions of vaccination.

**Table 5:** Caregiver perceptions on malaria vaccination.

| Variable |  | N (%) |
| --- | --- | --- |
| **Positive perception** | Vaccine is safe | 159 (39.1%) |
|  | Vaccine helps to prevent malaria | 284 (71.7%) |
|  | Vaccine is safe **AND** vaccine helps to prevent malaria | 114 (28.2%) |
|  | Vaccine is safe **OR** vaccine helps to prevent malaria | 325(81.8%) |
| **Negative perception** | The Vaccine is not efficient | 9 (2%) |
|  | The Vaccine can kill | 8 (1.9%) |
|  | The vaccine induces adverse effects | 13 (3.2%) |
|  | The Vaccine is not efficient **AND** The Vaccine can kill **AND** The vaccine induces adverse effects | 0(0) |
|  | The Vaccine is not efficient **OR** The Vaccine can kill **OR** The vaccine induces adverse effects | 23(5.7%) |
| **Mixed opinion** | Vaccine helps to prevent malaria **AND** The Vaccine is not efficient | 3(0.7%) |
|  | Vaccine helps to prevent malaria **AND** The vaccine induces adverse effects | 1(0.1) |

### 5. Comparison of the distribution of socio-demographic characteristics between vaccinated and unvaccinated children

Table 6 presents the distribution of sociodemographic characteristics between selected vaccination status. There was significant difference for sociodemographic characteristics between children with dose one malaria vaccination administration and those not benefiting dose one malaria vaccine administration. No significant differences were observed for caregiver age ≤19 years, occupation (farmer), religion (Muslim), or occupation (housekeeper) when comparing children who received the first dose on time versus those who did not, or for second-dose completeness.

**Table 6:** Sociodemographic characteristics distribution between selected vaccination status of children.

| Variables | Modalities | Dose 1 malaria administration |  | p-value |
| --- | --- | --- | --- | --- |
|  |  | Yes | No |  |
| **Cemented floor** | Yes | 74(37%) | 126(63%) | **<0.001** |
| **Caregiver being farmer** | Yes | 8(11.4%) | 62(88.6%) | **<0.001** |
| **Caregiver being Muslim** | Yes | 92(28.8%) | 227(71.2%) | **<0.001** |
| **Caregiver had Low level of education** | Yes | 32(21.6%) | 116(78.4%) | **<0.001** |
| **Caregiver being Mother** | Yes | 117(17.2%) | 256(82.8%) | **<0.001** |
| **Caregiver being female** | Yes | 119(30.7%) | 268(69.3%) | **<0.001** |
| **Caregiver being Housekeeper** | Yes | 61(30.7%) | 138(69.3%) | **<0.001** |
| **Caregiver being $\leq 19$ years** | Yes | 15(33.3%) | 30(66.7%) | **<0.001** |
|  |  | Dose 1 malaria vaccine Timeliness |  |  |
|  |  | Yes | No |  |
| **Cemented floor** | Yes | 38(51.4%) | 36(48.6) | **<0.001** |
| **Caregiver being farmer** | Yes | 4(49.4%) | 4(50.6%) | 0.057 |
| Caregiver being Muslim | Yes | 51(55%) | 41(45%) | 0.530 |
| Caregiver had Low level of education | Yes | 19(60.4%) | 13(39.6%) | <0.001 |
| Caregiver being Mother | Yes | 64(54.5%) | 53(45.5%) | <0.001 |
| Caregiver being Housekeeper | Yes | 34(55.5%) | 27(44.5%) | 0.742 |
| Caregiver being ≤ 19 years | Yes | 7 (44.8%) | 8(55.2%) | <0.001 |
|  |  | **Dose 2 malaria vaccine completeness** |  |  |
|  |  | Yes | No |  |
| Cemented floor | Yes | 47 (26.1) | 128 (73.9) | 0.001 |
| Caregiver being farmer | Yes | 4 (6.3) | 56 (93.7) | <0.001 |
| Caregiver being Muslim | Yes | 64 (22.3) | 214 (77.7) | <0.001 |
| Caregiver had Low level of education | Yes | 19 (15.2) | 105 (84.8) | <0.001 |
| Caregiver being Female | Yes | 77 (22.3) | 260 (79.7) | <0.001 |
| Caregiver being Mother | Yes | 75 (22.7) | 248 (77.3) | <0.001 |
| Caregiver being Housekeeper | Yes | 39 (23) | 127 (77) | 0.001 |
| Caregiver being ≤ 19 | Yes | 7 (20.3) | 27 (79.7) | 0.256 |

### 6. Association between caregivers’ perception and vaccination status of children

Table 7 presents the contribution of malaria vaccine perception on selected children vaccination status. Children exposure to caregivers with positive perception of malaria vaccination increased their chance to benefit the first dose malaria vaccination, to be administered this dose in time and to complete two doses malaria vaccination.

**Table 7:** Bivariate association between vaccine perception and vaccination status of children.

| Variables | Modalities | OR [95% CI] | P-Value | AOR [95% CI] | P-value |
| --- | --- | --- | --- | --- | --- |
| **Dose 1 malaria administration** |  |  |  |  |  |
| **Positive perception of malaria vaccine** | No (reference) |  |  |  |  |
|  | Yes | 12.66 [10.40-15.40] | **<0.001** | 15.54[12.63-19.10] | **<0.001** |
| **Dose 1 malaria vaccine timeliness** |  |  |  |  |  |
| **Positive perception of malaria vaccine** | No (reference) |  |  |  |  |
|  | Yes | 2.32[1.51-3.57] | **<0.001** | 2.36[1.52-3.65] | **<0.001** |
| **Dose 2 malaria vaccine completeness** |  |  |  |  |  |
| **Positive perception of malaria vaccine** | No (reference) |  |  |  |  |
|  | Yes | 8.42[6.75-10.50] | **<0.001** | 9.81[7.78-12.36] | **<0.001** |

\*cOR: Crude odds ratio; \*AOR: Adjusted Odds ratio; \*significant association cut-off: p<0.05

## Discussion

To the best of our knowledge, this is the first community-based survey exploring the coverage, completeness, and timeliness of malaria vaccination since its introduction in Cameroon. Results suggest that, documented vaccine coverage was 31.20% for dose 1, 22.61% for dose 2, and 17.70% for dose 3. The completeness of the second and third doses was 74.76% and 56.49%, respectively, and the timeliness of the first, second, and third doses was 55.24%, 50.35%, and 56.66%, respectively. Children exposure to caregivers with positive perception of malaria vaccination increased their chance to benefit the first dose malaria vaccination, to be administered this dose in time and to complete two doses malaria vaccination.

Vaccination coverage reflects the proportion of the target population that has benefited from the intervention. It is also an indicator of vaccine demand and of effectiveness of strategies implemented to reach targeted vaccination population. This study revealed that, less than one third of eligible children were documented to have benefited the first dose malaria vaccination. The second and third dose coverage of the malaria vaccination dropped to less than one fourth and to less than one fifth respectively. A study analysing administrative data reported a month following malaria vaccination introduction in Cameroon by the national EPI from the 42 targeted health districts revealed a first-dose malaria vaccine coverage relatively higher (37% of eligible children) than that of our study [24]. Fourteen months following the malaria vaccine introduction in Cameroon and one month prior to the data collected from this study, the three-dose malaria vaccination cumulative coverage from January to April 2025 reported by the national Expanded Program on Immunization was 64.3%, 54.1%, and 62.3% at national level, 53.9%, 40.6%, 42.2% in the two health districts targeted by our study for the first, second and third malaria vaccination dose respectively [25]. The discrepancies between these administrative data and this study is yet to be understood and can be explained by the children reached during vaccination session are not rigorously selected from communities belonging to vaccinating health district, by the fact that our study estimated only documented vaccination, and by probable error when estimating the numerator or denominator of administrative malaria vaccination coverage. The relatively higher malaria vaccination coverage estimated after the vaccine introduction in other countries can be explained by better communication prior to the vaccination introduction, resulting in better adherence of the caregivers and/or by vaccination strategies [16, 26]. The coverage estimated from our study was below expectation of the national Cameroon EPI [27]. Assuming that the malaria vaccine introduced in Cameroon (RTS,S/AS01) retains its efficacy as reported in clinical trials, the three-dose vaccination coverage reported in our study predicts that nearly one in twenty children reaching their first birthday will benefit from the vaccine’s protection against malaria in the two targeted districts. This underlines the urgent need, on the one hand, to investigate the determinants of poor performance of malaria vaccination and, on the other hand, to implement documented interventions in the same context that has been shown to contribute to improving EPI vaccination coverage [23].

The vaccination completeness indicator is important for monitoring the performance of immunization programs [28]. It also provides information on the health system’s ability to ensure optimal protection for vaccinated children, as vaccine effectiveness depends on full compliance with the recommended vaccination schedule for each given vaccine. Malaria vaccination completeness is necessary to maximize individual immunity but also to significantly reduce malaria morbidity and mortality in children [11]. In this study, half of the children who started the malaria vaccine continued through to the third dose. The observed high dropout rate in our study appears to be consistent with varying levels of importance for malaria vaccine and other EPI vaccines as documented from studies conducted in similar and other contexts [29]. The present study was not planned to understand factors catalysing malaria vaccination dropout rate. In the absence of access to studies exploring vaccine completeness for malaria vaccination, the consultation of studies exploring it for other vaccines identified negative maternal perceptions of vaccination, the birth of the child in the community and limited geographical accessibility to vaccination appear to increase the risk of vaccine incompleteness [30]. Further studies should explore the contribution of these factors on malaria vaccination completeness and others such as the scheduling of vaccination sessions and the occurrence of adverse events after vaccination identified to induce vaccination hesitancy [31]. The benefit of other caregivers reminders using SMS or, tracking of children vaccination status in households by community volunteers to catch up on missed doses, shown to be beneficial for other vaccines, should be explored for their contribution to improving malaria vaccination completeness [23, 31].

The timing and recommended interval between vaccine doses are based on the immune response and child development, as assessed through clinical trials and age-specific disease risks [32, 33]. To our knowledge, no studies have yet been conducted to assess the timeliness of malaria vaccination in Cameroon. Like all EPI vaccines, malaria vaccination has an administration schedule, but this schedule does not provide an operational definition of vaccination timeliness. Considering in our study an administration of each dose of malaria vaccine within one month following the exact age of administration recommended in the EPI vaccination schedule, it turned out that approximately half of the children received their dose of vaccine on time. Comparing this trend with the timeliness of other EPI vaccines administered in the same context a few years earlier shows a higher timeliness rate for the BCG vaccine, which is administered at birth, and a timeliness rate consistent with our study for the DPT-Hi + Hb 3 vaccine, i.e., nearly half of children receiving the third dose of this vaccine within one month of the age recommended by the EPI [29]. This consistency probably matches the fact that nearly half of vaccinated children are vaccinated later than recommended is related to the supply or demand of all EPI vaccines and not just the malaria vaccine and has to be addressed by the program. Follow-up studies of children in high-risk areas show an increase in malaria incidence with increasing time following the administration each malaria vaccine dose, suggesting that some of the benefit of malaria vaccine is lost when the vaccine administration schedule is not respected [11]. Making available an operational definition of the timeliness of each vaccine dose, conducting studies to understand low vaccine timeliness, and identifying interventions to improve it should guide the Expanded Program on Immunization to improve malaria vaccine timeliness.

Caregivers’ perception of children vaccination is expected to influence it demand in the benefit of children. In this study, a positive perception of the malaria vaccine was documented to significantly increase children access to the first malaria dose as well as the timeliness of the dose and the completeness of the second dose. To the best of our knowledge no published study has investigated the contribution of caregiver perception on malaria vaccine and children access to this vaccine. In Malawi, revealed evidence in line with our finding by documenting the contribution of caregivers’ exposure to negative rumors on the malaria vaccine and significant reduction of children likelihood of full uptake of the malaria vaccine [34]. This finding goes beyond the scope of the malaria vaccine. Indeed, a systematic review of general childhood vaccination revealed that negative perceptions of vaccines were significantly associated with non-use of vaccination [35]. Therefore, generating evidence on innovative interventions to improve caregivers’ perception on malaria vaccine is expected to guide decision makers in taking needed action to improve malaria vaccine uptake in children.

The results of this study should be used taking in consideration following limitation. Malaria vaccination coverage, timeliness and completeness were estimated based on vaccination status. This could underestimate access to vaccination, as a given proportion of vaccinated children may be vaccinated without documentation [36].

## Conclusion

In conclusion, the coverage, completeness, and timeliness of malaria vaccination are below the targets needed to achieve optimal vaccination benefits. Children exposure to caregivers with positive perception of malaria vaccination increased their chance to benefit the first dose malaria vaccination, to be administered this dose in time and to complete two doses malaria vaccination. We recommend that the competent health authorities in the targeted health districts to implement communication initiatives aimed at improving caregivers’ perceptions on malaria vaccination; that scientists test innovative interventions to improve caregivers’ perceptions on malaria vaccination and that other determinants of children’s access to malaria vaccination be explored to guide decision-making on promoting children’s access to the malaria vaccine.

## Data Availability

The database is available and accessible from the corresponding author.

## Acknowledgments

We thank the health authorities of the Rest Region of Cameroon for facilitating and accomodating the implementation of the project. We also thank all enumerators and supervisors for their contribution in the data collection.

